# Estimating age patterns and grouped temporal trends in human contact patterns with Bayesian P-splines

**DOI:** 10.64898/2026.01.21.26344589

**Authors:** Bryan Sumalinab, Oswaldo Gressani, Niel Hens, Christel Faes

## Abstract

This paper presents a smoothing method to estimate age-specific human contact patterns and their variations over different periods. Specifically, it examines how age-specific contact patterns shift under varying conditions, such as holiday periods and levels of public health intervention. The method uses Bayesian P-splines to smooth age-specific contact rates and leverages Laplace approximations for fast Bayesian inference, significantly reducing computational complexity. The proposed methodology is applied to the CoMix data from Belgium, a social contact survey collected during the COVID-19 pandemic. Results indicate significantly reduced contacts during periods in which strict social policies were in place, particularly among adults, and notable reductions among young individuals during holidays. This research advances our understanding of how human contact adapts in response to varying social and policy conditions, which can guide more realistic and adaptive infectious disease transmission models.

## 1 Introduction

The transmission of respiratory infectious diseases, such as COVID-19 and influenza-like illnesses, is traditionally driven by human interactions, where pathogens spread through direct or indirect contact (Randall et al., 2021). Understanding how people interact within different social and demographic contexts is, therefore, crucial to mitigate the spread of such diseases and accurately describe disease transmission (Prem et al., 2020; Hoang et al., 2022). Epidemiologists rely on mathematical models to capture these interactions, estimate transmission parameters, investigate the impact of control strategies, and predict outbreak trajectories (Wallinga et al., 2006; Mossong et al., 2008; Hens et al., 2009; Fumanelli et al., 2012). Mathematical models have long been used to understand and control the spread of infectious diseases. One of the key parameters of mathematical models is the transmission rate, which depends on the mixing behavior of individuals within a population. Quantifying these mixing patterns requires empirical data, often collected through social contact surveys. A review of such surveys can be found in Hoang et al. (2019) and Mousa et al. (2021).

Studies have shown that age is one of the most influential factors affecting contact patterns and, consequently, the transmission of infectious diseases (Hoang et al., 2019; Wallinga et al., 2006). For instance, children tend to have higher contact rates, whereas older adults generally engage in fewer social interactions. Additionally, individuals are more likely to interact with others of similar age groups, a phenomenon known as age-assortative mixing (Hens et al., 2009; Mossong et al., 2008). Contact patterns also vary by time and setting. A review of social contact studies found that the number of contacts is generally higher during school terms compared to school holidays. Similarly, notable differences have been observed between weekdays and weekends, with individuals typically reporting more contacts on weekdays (Hoang et al., 2019). During the COVID-19 pandemic, non-pharmaceutical interventions (NPIs) such as lockdowns and physical distancing measures were also shown to significantly reduce the number of social contacts (Liu et al., 2021).

These factors can influence not only the overall number of contacts but also the patterns of age-specific mixing, that is, interactions between individuals of different age groups. This raises an important research question: how do age-specific contact patterns shift under varying conditions, such as holidays versus term time, weekdays versus weekends, and in response to public health interventions? Understanding these variations is crucial for assessing the spread of infectious diseases, optimizing the timing of interventions, and evaluating the effectiveness of control strategies. Although several methods have been proposed for estimating age-specific contact rates (Mossong et al., 2008; Hens et al., 2009; Goeyvaerts et al., 2010; van de Kassteele et al., 2017; Vandendijck et al., 2024), these approaches are often limited in their ability to capture changes in contact patterns across different contexts. This challenge is further complicated by the interaction between contact rates and the fabove-mentioned factors, which introduces an additional layer of complexity. Moreover, the reciprocal nature of contacts, which will be discussed in the next section, imposes constraints that standard smoothing methodologies cannot accommodate. The larger volume of data involved also implies computational challenges. In this paper, a new modeling strategy is proposed to address these issues. We propose a negative binomial model with overdispersion parameter modeled as a two-dimensional function of the participant’s age and contact’s age. This permits to capture unmeasured age-specific variability and potentially leads to a more accurate estimation of uncertainty.

Our method builds upon the Laplacian-P-splines (LPS) framework, which has been successfully applied in various epidemiological contexts, such as nowcasting (Sumalinab et al., 2025, 2024), estimation of the reproduction number (Gressani et al., 2022, 2023), and estimation of incubation periods (Gressani et al., 2025). The key idea is to smooth age-specific contact rates using Bayesian P-splines, defined on a one-year age interval, and to approximate the conditional posterior distribution of the latent parameters by means of Laplace approximations. LPS offer a flexible framework for modeling age-specific contact rates in a fully Bayesian setting with a natural quantification of uncertainty. The Laplace approximation opens up a sampling-free approach to inference and permits to bypass computationally intensive Markov chain Monte Carlo (MCMC) methods. This advantage is particularly useful given the complexity of the model and the large amount of data underpinning in our analysis.

The paper is organized as follows. Section 2 presents the model for age-specific contact rates with an age-varying overdispersion parameter and the implementation of a reciprocity constraint. We also describe the penalization approach used to enforce this constraint and explain how the penalty structure translates into priors for the penalty parameters. Additionally, we discuss the posterior distributions and the use of Laplace approximations to estimate the conditional posterior distribution of the latent parameters. In Section 3, the methodology is applied to data from the CoMix study in Belgium by considering two grouping strategies: the first approach groups the survey waves according to the policy stringency index, while the second approach groups them by holiday versus non-holiday periods. Changes in age-specific contact rates are then examined within each group. Finally, Section 4 concludes the paper with a summary and discussion. The code used to reproduce the results of this article is publicly available at https://github.com/bryansumalinab/Social-Contact-Smoothing—LPS.

## 2 Methods

### 2.1 Contact rate model

Let *Y*_*ijt*_ denote the total number of individuals of age *j* contacted by participants of age *i* for *i* = 1, …, *m* and *j* = 1, …, *m* at time *t* = 1, …, *T* (the time point when the survey is conducted). Denote the mean number of individuals of age *j* contacted by a participant of age *i* at time *t* by *m*_*ijt*_ = 𝔼(*Y*_*ijt*_)*/r*_*it*_ where *r*_*it*_ is the total number of participants of age *i* at time *t*. The per capita contact rate is given by *c*_*ijt*_ = *m*_*ijt*_*/p*_*jt*_, where *p*_*jt*_ is the total population of age *j* at time *t*.

We assume a negative binomial distribution for *Y*_*ijt*_ with mean 𝔼(*Y*_*ijt*_) = *µ*_*ijt*_ and variance 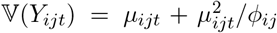, where the overdispersion parameter *ϕ*_*ij*_ is age-specific. Let *g* ∈ {1, 2, …, *G*} denote a categorical group variable. For example, *g* = 1 could represent a collection of different surveys conducted during periods when the policy stringency index is high. Similarly, *g* = 2 may represent surveys from periods with moderate stringency, and so on for additional group levels. Note that the grouping variable *g* may also be defined at the time point level, in which case each time *t* is uniquely associated with a group, i.e., *g* = *t*. Suppose we consider group *g* = 1 to be the reference category and define an indicator function:

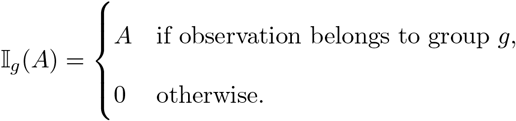

This allows us to formulate a mean regression model given by:

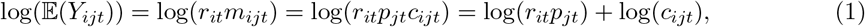

and offsets log(*r*_*it*_*p*_*jt*_) where the per capita rate is modeled as a smooth function

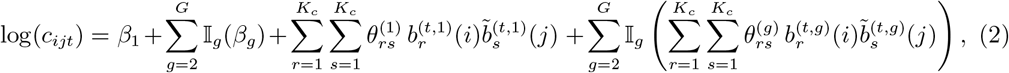

where 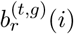 and 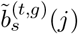 are cubic B-spline basis functions for the age of participants and age of contacts, respectively, for group *g* at time *t, K*_*c*_ is the number of basis functions, and 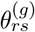 are the B-spline coefficients for each group. For the overdispersion parameter, an age-specific two-dimensional model is assumed:

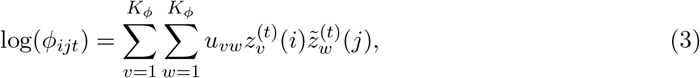

where 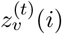 and 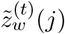 are B-spline basis functions for the age of participants and age of contacts at time *t* with corresponding coefficients *u*_*vw*_, and *K*_*ϕ*_ is the number of basis functions for the model of the overdispersion parameter.

An important constraint for the contact rates is the reciprocity requirement. This means that, at the population level, the total number of contacts between individuals of age *i* with individuals of age *j* must be equal to the total number of contacts from age *j* to *i*, i.e., *m*_*ijt*_*p*_*it*_ = *m*_*jit*_*p*_*jt*_ for each *t*, or equivalently, *c*_*ijt*_ = *c*_*jit*_. To illustrate how the reciprocity constraint is imposed in our model, we consider only group 1 (i.e., *g* = 1) for the contact rate model in (2) given by:

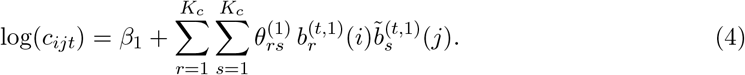

Each *c*_*ijt*_ in (4) involves the expression

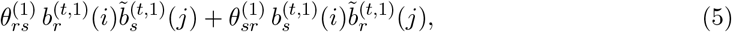

while the corresponding *c*_*jit*_ term involves

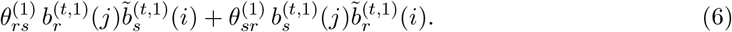

If we assume symmetry in contact rates, i.e., *c*_*ijt*_ = *c*_*jit*_, then equations (5) and (6) must be equal. This holds only if 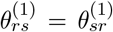. The same reciprocity condition applies to the B-spline model for other groupings of periods. Hence, in general, imposing the constraint 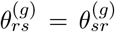 ensures symmetry in the contact rate matrix, and thus satisfies the reciprocity constraint.

### 2.2 Penalization and prior distributions

Let ***θ***_*g*_ denote the vector of B-spline coefficients obtained by stacking the columns of the full *K*_*c*_ *× K*_*c*_ spline coefficient matrix **Θ**_*g*_ for group *g* given by:

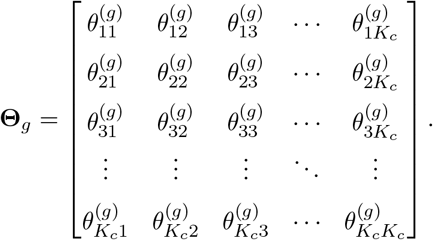

In the context of P-splines (Eilers and Marx, 1996), a discrete difference penalty is imposed on successive B-spline coefficients. The penalty is translated to a Gaussian prior in a Bayesian context (Lang and Brezger, 2004), given by 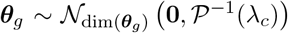, where *λ*_*c*_ is a positive smoothing parameter that controls the roughness of the fitted contact rate surface. The precision matrix is defined as 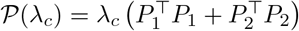 with 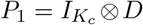 and 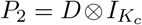 representing the row-wise and column-wise difference operators, respectively, applied to the matrix Θ_*g*_. Here, *D* is the one-dimensional difference matrix. Due to the symmetry constraint for Θ_*g*_, it is sufficient to estimate only its lower triangular part, including the main diagonal. Consequently, the penalty is applied only to the row-wise and column-wise differences corresponding to these elements, similar to the penalty used by van de Kassteele et al. (2017). Furthermore, the precision matrix 𝒫 (*λ*_*c*_) is rank deficient, and this issue is typically addressed by adding a small positive constant to its main diagonal to ensure full rank and numerical stability. For the overdispersion model in (3), the penalty matrix is constructed similarly but without the symmetry constraint. Thus, the prior for the vector of coefficients ***u*** is given by ***u*** ∼ 𝒩_dim(***u***)_ **0**, 𝒫^−1^(*λ*_*ϕ*_) where *λ*_*ϕ*_ is the penalty parameter for the fitted overdispersion surface.

An uninformative Gaussian prior is assumed for the parameters ***β*** = (*β*_1_, *β*_2_, …, *β*_*G*_)^⊤^ in (2), namely 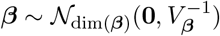 with *V*_***β***_ = *ζI*_*G*_ (*ζ* = 10^−5^), where *I*_*G*_ denotes a *G × G* identity matrix. Following Jullion and Lambert (2007), we impose robust priors on the penalty parameters given by 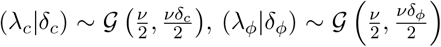 and *δ*_*c*_ ∼ 𝒢 (*a*_*δ*_, *b*_*δ*_), and *δ*_*ϕ*_ ∼ 𝒢 (*a*_*δ*_, *b*_*δ*_), with *a*_*δ*_ = *b*_*δ*_ chosen to be sufficiently small (e.g., 10^−5^) with fixed *ν* = 3. 𝒢 (*a, b*) denotes a Gamma distribution with mean *a/b* and variance *a/b*^2^. Define the precision matrix for all B-spline coefficients of the contact rate model by 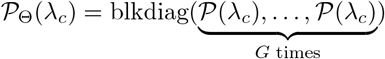, the latent parameter vector ***ξ*** = (***β, θ***_1_, ***θ***_2_, …, ***θ***_*G*_, ***u***)^⊤^ and the precision matrix 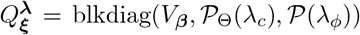, where blkdiag(·) denotes a block diagonal matrix. The prior for ***ξ*** conditional on the penalty vector ***λ*** = (*λ*_*c*_, *λ*_*ϕ*_)^⊤^ is then written as 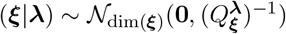.

### 2.3 Posterior distributions and Laplace approximation

We approximate the conditional posterior distribution of the latent parameters ***ξ*** given ***λ*** using the Laplace approximation, following the LPS methodology of Gressani and Lambert (2018, 2021). This results in a Gaussian approximation centered at the conditional posterior mode 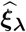, with covariance matrix 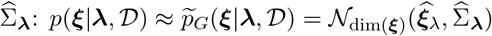. The penalty parameters are fixed at their maximum a posteriori values.

## 3 Data application

### 3.1 Data

We illustrate our method using the CoMix data from Belgium, which was collected through a longitudinal survey conducted during the COVID-19 pandemic. The survey gathered data from a representative sample of the Belgian population based on age, gender, and region to ensure demographic balance. Data collection occurred bi-weekly between April 2020 and August 2020, and again from November 2020 to March 2022, resulting in a total of 43 survey waves (Coletti et al., 2020; Loedy et al., 2023). Participants provided detailed daily records of their social contacts, with each contact defined as either a physical interaction or an in-person conversation involving at least a few words. The CoMix data spans different holidays and multiple phases of public health interventions (including mainly NPIs), ranging from strict lockdowns to more relaxed periods, offering valuable insights into how social contact patterns shifted during holiday periods and in response to evolving policy measures. Coletti et al. (2020) and Loedy et al. (2023) give more details about NPIs implemented in Belgium during the survey period.

Our analysis only includes waves 12 to 43. Children did not participate in the first eight waves, and there were changes in survey design following wave 11, making earlier data less consistent. Furthermore, the age of the participants under 18 years old is reported in categories ([Under 1], [1–4], [7–11], [12–15], [16–17]), typically filled in by parents. We assign participants’ ages within these groups by sampling uniformly within each category. Similarly, only the age range of their contacts was reported for some participants. In such cases, we also sample ages uniformly within the reported contact age ranges. Population counts for each age in the year 2021 were obtained from Statbel (Statbel, 2021).

### 3.2 Results

In the analysis, we considered two different grouping strategies. First, we categorized the survey according to the policy stringency index (low, medium, high). The stringency index summarizes various government measures implemented in response to COVID-19. This index ranges from 0 to 100, where 0 indicates no intervention and 100 reflects the most stringent measures, with data obtained from Hale et al. (2021). We grouped the survey periods in three classes according to low, medium and high stringency. Second, we grouped the data based on whether the survey period overlapped with a major holiday, having two categories: holiday and non-holiday waves. The waves classified as holiday periods include Wave 12 (Christmas 2020), Wave 20 (Easter 2021), Waves 26–29 (Summer 2021), and Wave 38 (Christmas 2021). Figure 1 shows the different waves considered in the analysis, including the start date of the survey and the average stringency index for each wave. Each grouping strategy is analyzed using two models, one with group interaction terms and one without. The model without interaction assumes that changes in age-specific log-contact rates vary only by an additive constant across groups. That is, the shape of the contact pattern remains the same across groups, with only overall levels shifting. In contrast, the model with interaction (i.e., equation (2)) allows the changes in log-contact rates to vary differently for different ages across different groups, capturing more complex group-age-specific dynamics. We compared the models using the Watanabe-Akaike Information Criterion (WAIC) (Watanabe, 2013), also referred to as the Widely Applicable Information Criterion, where lower values indicate a better model fit. For both grouping strategies, the models that incorporate group interactions exhibit substantially lower WAIC values compared to those without interactions (see Table 1), indicating that changes in log-contact rates vary by age across different groups. Therefore, we focus on the results obtained from the models including interaction terms.

**Table 1.** WAIC values for models grouped by holiday periods and stringency index, fitted with and without interaction terms.

| Groupings | Model | WAIC |
| --- | --- | --- |
| Holiday | without interaction | 205811.50 |
|  | with interaction | 204990.10 |
| Stringency | without interaction | 205649.30 |
|  | with interaction | 204419.60 |

**Figure 1.**
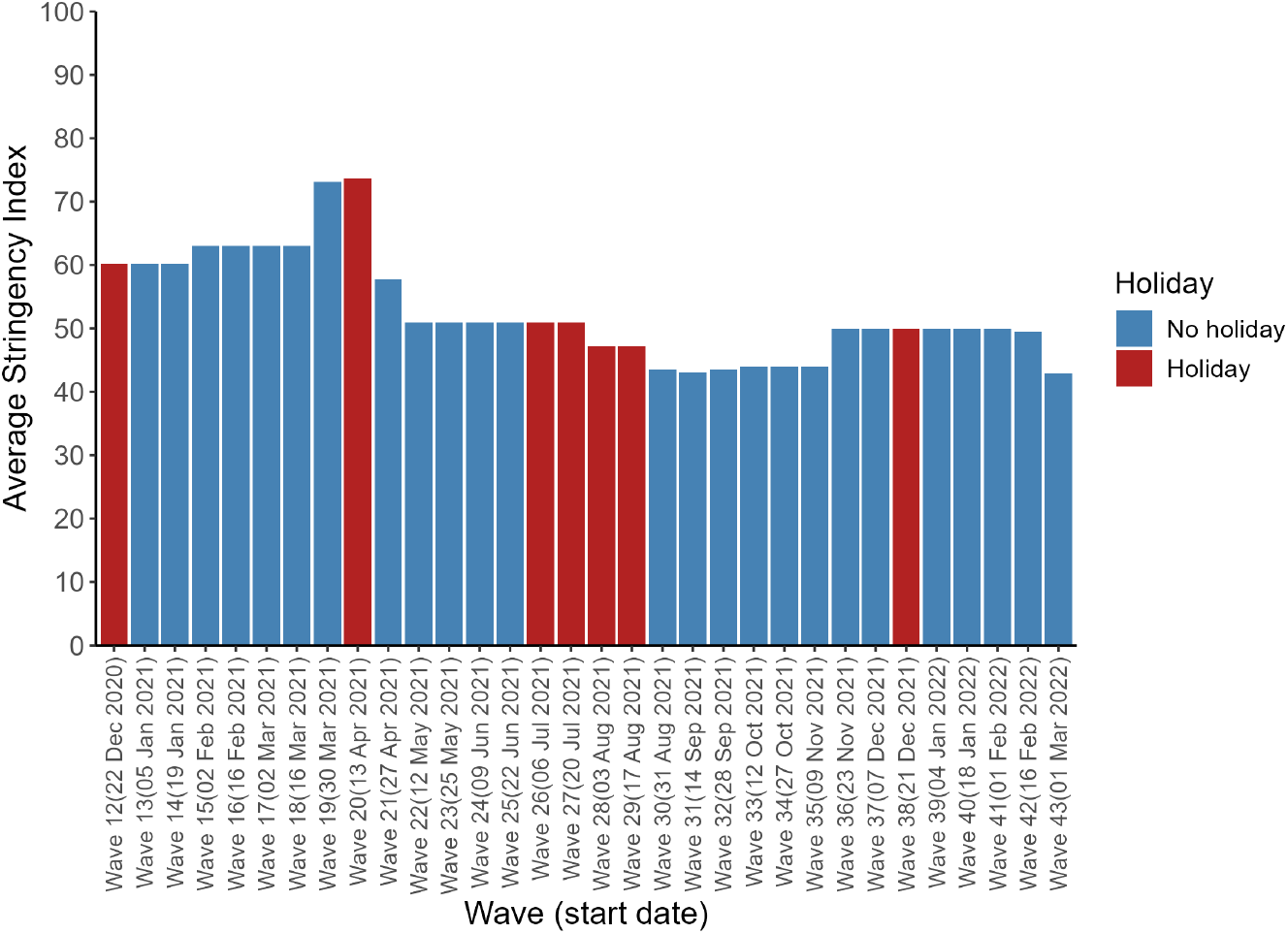
Stringency index during the survey time period. *High stringency* waves 12 to 21; *Moderate stringency* - waves 22 to 29 and 36 to 42; *Low stringency* - waves 30 to 35 and 43

Moreover, the difference in estimated contact rates relative to the reference category is computed to determine which age groups show significant differences. For example, for the stringency index grouping, the reference category is low stringency, and differences are calculated between moderate and low stringency, as well as high and low stringency levels. Additionally, high and moderate stringency levels are also compared. To assess the significance of these differences, note that from the Laplace approximation scheme, the log contact rates are approximately Gaussian distributed. Thus, we sample 1000 times from this Gaussian distribution to compute the differences, and obtain a 95% quantile-based credible interval by calculating the 2.5% and 97.5% quantiles of the approximate posterior distribution of differences. This interval allows us to test the null hypothesis that the difference is zero. If zero is not included in the interval, we interpret the difference as statistically significant at a 5% significance level.

Figure 2 shows the estimated log-contact rates for each group of the stringency index. The overall pattern follows the typical shape of age-specific contact rates, with younger individuals generally having the highest contact rates. Figure 3 shows how different levels of policy stringency affect age-specific contact rates. Both moderate and high stringency generally lead to fewer contacts compared with low stringency, indicated by large parts of negative values of the difference (Figure 3a). Comparing high stringency and low stringency (Diff_HL_), most reductions during high stringency occur among people aged 40–80 interacting with others in the same age range. Figure 3b confirms that most of these negative values are statistically significant. Additionally, fewer contacts are observed between younger individuals and adults under high stringency. A few positive values, indicating higher contact rates under strict measures, appear primarily among younger groups, with only a few of these differences being statistically significant (see Diff_HL_ in Figure 3b). On the other hand, moderate stringency (Diff_ML_) produces a more varied direction of the difference and weaker effects. Figure 3a shows mixed positive and negative regions with a lower overall magnitude of the difference (Figure 3c). Only very few positive differences are significant, notably among individuals around 60 years old, while statistically significant negative differences mainly involve children-to-children and children-to-adult contacts (Figure 3b). Moreover, the comparison between high and moderate stringency (Diff_HM_) shows reductions in contact rates under high stringency among older participants. These reductions are statistically significant, suggesting that stricter NPIs lead to further reductions in contact rates compared to moderate measures. In contrast, younger participants tend to have higher contact rates under the most stringent policies compared to moderate or less strict measures. We additionally fitted the same model restricted to non-holiday wave periods only, and the results are generally similar (see Supplementary Material).

**Figure 2.**
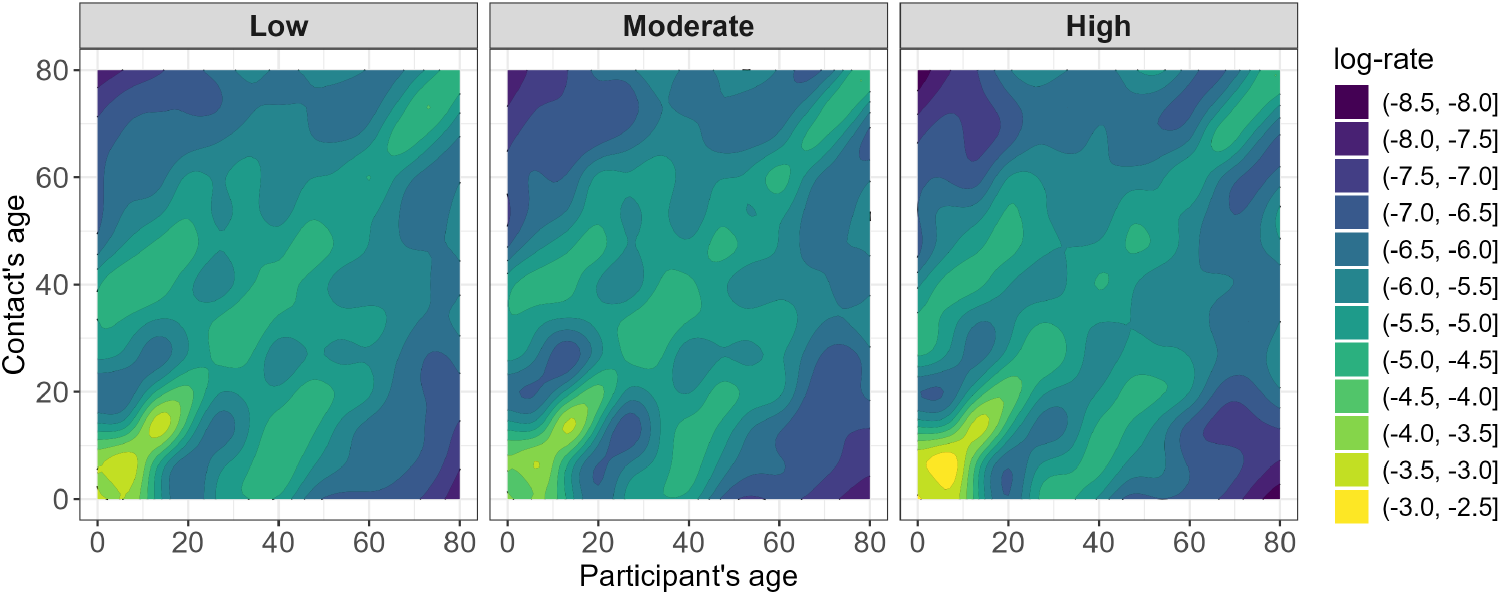
Estimated log-contact rates per 10,000 during periods with low, moderate or high stringency measures.

**Figure 3.**
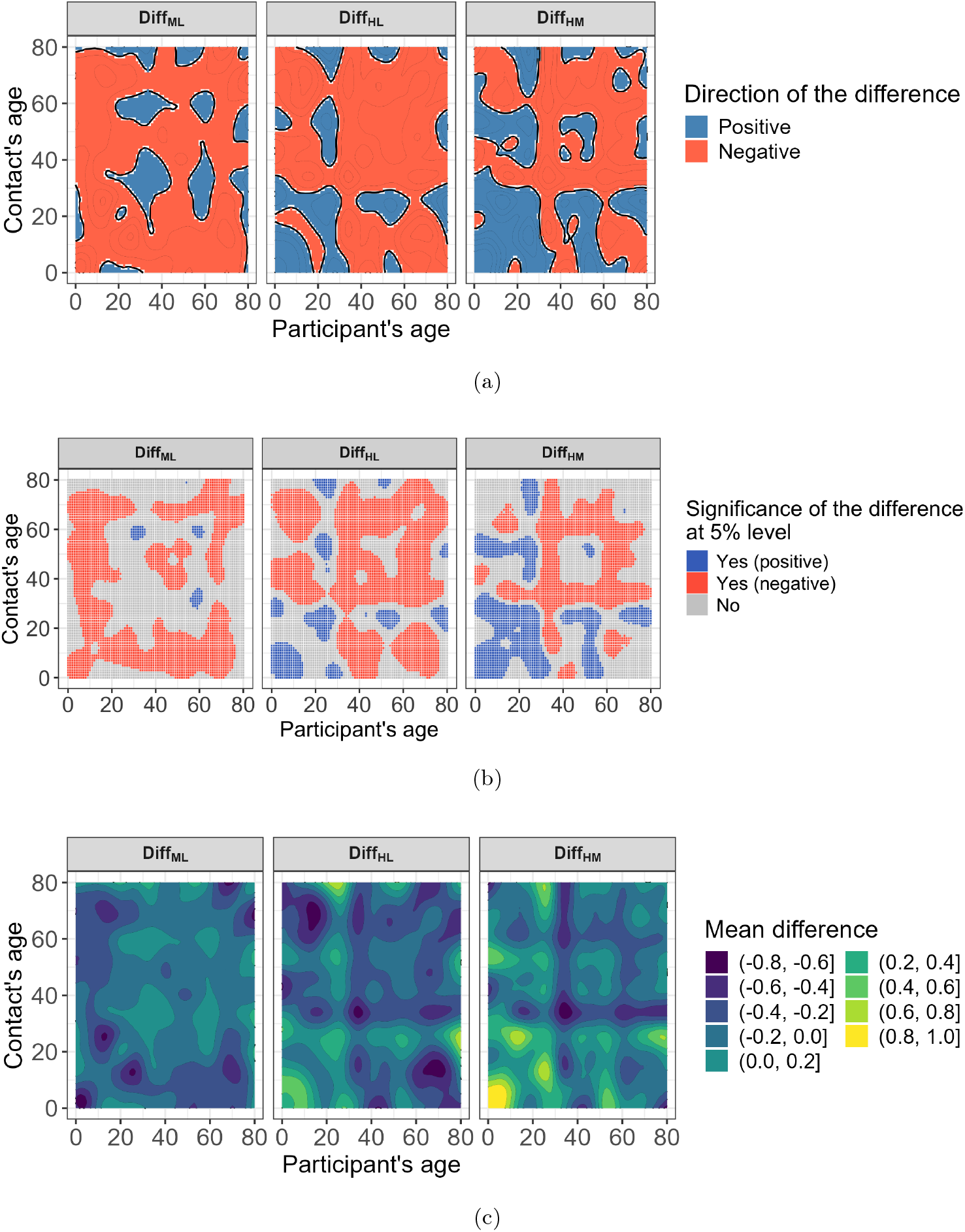
Differences in contact rates between different stringency indices, based on the model with stringency interaction. *Diff*_*ML*_ = Moderate - Low Stringency; *Diff*_*HL*_ = High - Low Stringency; *Diff*_*HM*_ = High - Moderate Stringency. (a) Direction of the difference (*Negative* = fewer contacts compared to low stringency index; *Positive* = more contacts com- pared to low stringency index); (b) Statistical significance of the difference at a 5% significance level (*Yes (positive)* = Significant and positive direction of the difference; *Yes (negative)* = Significant and negative direction of the difference; *No* = Not significant); (c) Mean difference.

Regarding holiday effects, the estimated contact patterns are presented in Figure 4. Decreases in contact rates during holidays (i.e., negative differences in Figure 5a) are most pronounced in interactions involving younger individuals (approximately ages 0–25 years), particularly with peers or adults aged 25–60 years. Statistically significant reductions (Figure 5b) are concentrated in youth-to-youth interactions (ages 0–20 years), as well as between children (ages 0–10 years) and middle-aged adults (ages 40–60 years), and between adolescents (ages 10–20 years) and working-age adults (ages 20–60 years). Notably, significant declines are also observed in interactions within middle-aged adults (around age 50 years) and among older adults (ages 60–80 years). The largest negative differences (Figure 5c) occur among young individuals (ages 0–20 years) interacting primarily with others in the same age group. On the other hand, significant increases in contact rates during holidays are observed among adults aged 30–40 years interacting with peers in the same age group, as well as older adults (around age 60) interacting broadly across a wide age range (20–80 years). Additionally, a notable increase occurs between older adults aged 70–80 years and those aged 50–60 years. The most significant increase occurs in interactions between adults around age 60 years and middle-aged individuals (around 40 years old) or the oldest group (around 80 years old). Finally, Figure 6 presents the smooth age-varying overdispersion parameter for the two models with group interaction. Regions with higher overdispersion correspond to areas in the data with generally more frequent zero values, which may contribute to the higher estimated overdispersion.

**Figure 4.**
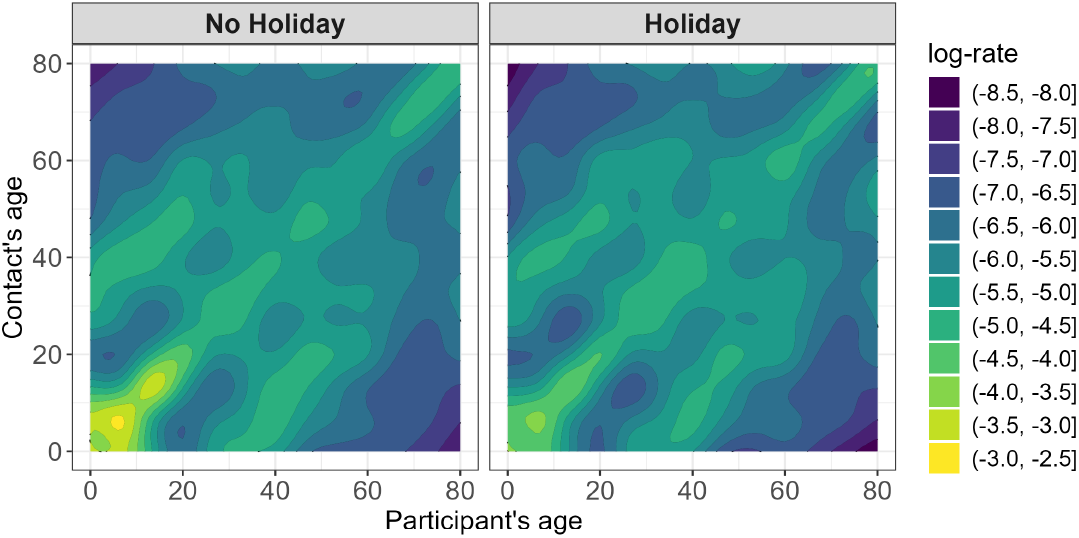
Estimated log-contact rates per 10,000 based on the model with holiday interaction.

**Figure 5.**
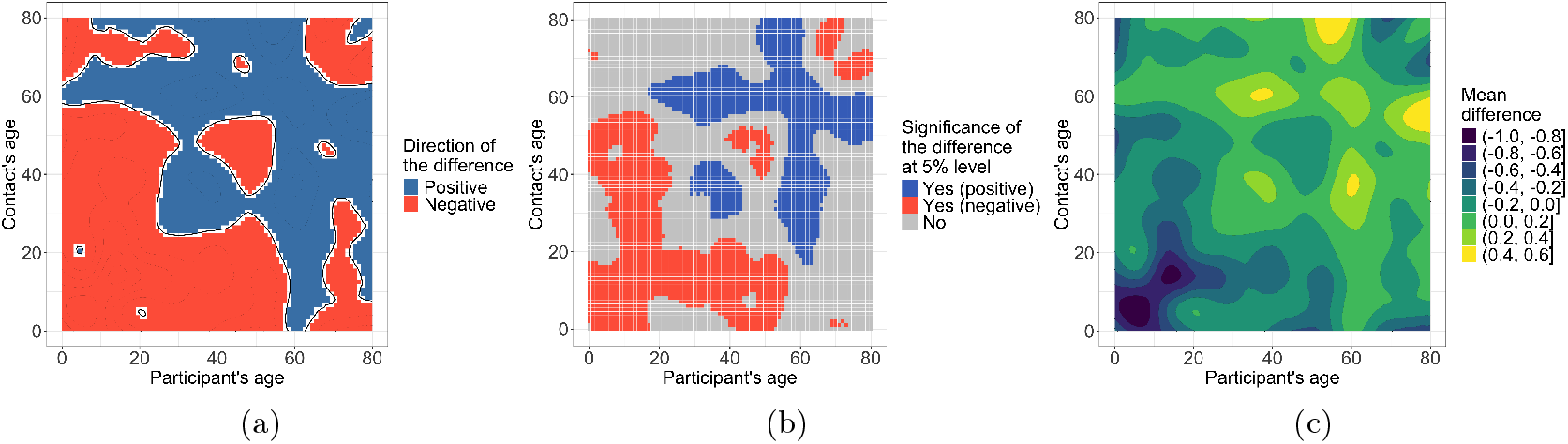
Differences in contact rates between holidays and non-holidays, based on the model with holiday interaction (*Difference* = Holiday – No Holiday). (a) Direction of the difference (*Positive* = more contacts on holidays; *Negative* = fewer contacts on holidays); (b) Statistical significance of the difference at a 5% significance level (*Yes (positive)* = Significant and positive direction of the difference; *Yes (negative)* = Significant and negative direction of the difference; *No* = Not significant); (c) Mean difference.

**Figure 6.**
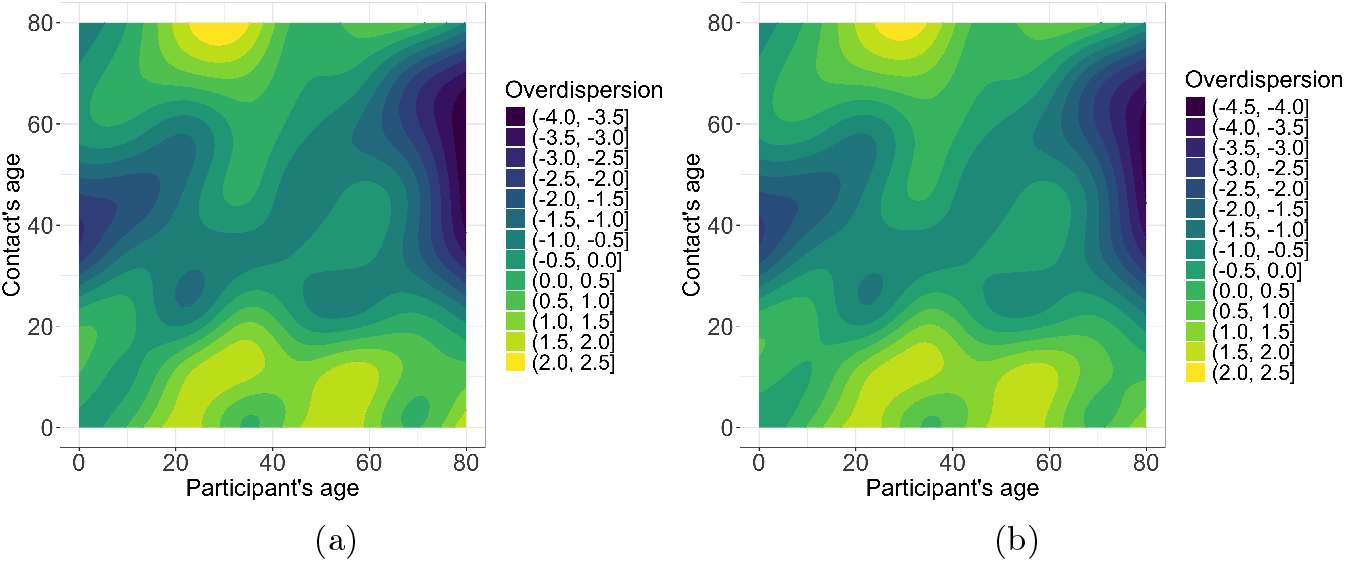
Estimated age-varying log-overdispersion 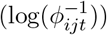 (a) Model with holiday interaction; (b) Model with group interaction based on stringency index groupings.

## 4 Conclusion

This study introduces a method to better understand how people of different ages interact by looking at factors like holidays and policy strictness that modify the social contacts of individuals across different age groups. Investigating such factors is important because they can significantly influence the transmission dynamics of infectious diseases, inform targeted public health interventions, and improve the accuracy of epidemic modeling. The LPS approach enables fast and flexible Bayesian estimation of contact rates by accounting for contact reciprocity and age-varying overdispersion. As a sampling-free method, it avoids reliance on MCMC sampling, which can be computationally demanding given the complexity of the considered model class. Our method is beneficial not only for investigating temporally grouped effects but also for estimating age-specific social contact rates by combining multiple surveys conducted over comparable periods. This approach makes a better use of available data to maximize information while remaining computationally efficient.

The data application shows how the level of policy stringency and holidays affect age-specific social mixing patterns. The analysis of the stringency index indicates that higher levels of policy stringency are generally linked to reduced social contacts, consistent with previous empirical findings (Liu et al., 2021). Notably, we observed fewer adult interactions during periods of strict measures compared to times with less stringent restrictions. This likely reflects an increase in adult contacts during post-relaxation periods that showed an increase in contact rates primarily among working-age adults following the reopening of workplaces (Liu et al., 2021). Conversely, younger individuals tended to have more contacts during periods of high stringency index compared to those with less strict measures. With regards to holidays, we observed the largest and most significant reductions in social contact among younger individuals during holidays, likely reflecting the drop in school-related interactions (Eames et al., 2011; Chen and You, 2015). Interestingly, our findings also suggest that children and adolescents have fewer interactions with adults during holidays, contrary to the results of Eames et al. (2011). Moreover, adults aged approximately 30–40 showed an increase in contacts with individuals of similar age during holidays, a pattern that aligns with findings of Hens et al. (2009) for Belgium (though in their study, among 25–35 year-olds). This may be due to social gatherings or holiday-related events among adults in this age group. We also observed an increase in intergenerational mixing during holiday periods. However, in our analysis, this increase was primarily among adults rather than between children and (grand) parents. Specifically, individuals around 60 years old have more interactions with younger, middle-aged, and older adults within the 20–80 age range during holidays.

Although our findings are largely consistent with previous literature and offer valuable insights, they should be interpreted with caution due to certain limitations. For example, the survey was not explicitly designed to examine the effects of holidays on social contact patterns. Since the data were collected during the COVID-19 pandemic, the level of policy stringency may confound the observed effects of holidays, and vice versa. In particular, some holiday periods coincided with different stringency phases, making it difficult to disentangle their individual contributions. Furthermore, because each survey wave spans two weeks, the results may be influenced by weekend effects. In addition, some minor holidays were included as regular periods, as our analysis focused only on major holidays. Moreover, to fully understand the context of these changes in social contact patterns, further investigation is needed, particularly into where these contacts occurred (e.g., at school, in the workplace, or at home). Regarding the sampling scheme for the reported contact age intervals, it may be useful to consider sampling within narrower age intervals in certain settings, particularly when the reported age range is wide. For example, in school settings, children are more likely to have contacts with peers close in age (e.g., within ±1 year) than those significantly older or younger.

Another limitation is that we did not account for the longitudinal structure of the data, where the same participants took part in successive survey waves. Incorporating this repeated-measures structure could improve model accuracy and precision by accounting for within-person correlations over time. This points to an additional issue, a potential under-reporting due to fatigue. Participants who respond over multiple waves may gradually underreport their contacts, which could introduce a downward bias in the reported number of contacts. Accounting for such effects could further enhance the reliability of the model estimates (Loedy et al., 2024; Dan et al., 2024). In the analysis, we grouped the survey waves into three levels of policy stringency: low, medium, and high stringency. However, a more granular analysis could be performed by treating each wave individually. We explored this approach, but encountered numerical issues due to the increased complexity. An alternative approach could involve treating survey waves as a continuous temporal surface and modeling the interaction between age and time using a three-dimensional smoothing model. This would extend the current two-dimensional framework to incorporate time as a third dimension. Finally, future work could also extend the Laplacian-P-splines framework to estimate age-specific contact rates stratified by gender and potentially investigate gender-specific changes across grouped temporal trends. A key challenge in such an extension is that it requires a more tailored implementation of the reciprocity constraint.

## Data Availability

All data produced are available online at https://socialcontactdata.org/data/.

https://github.com/bryansumalinab/Social-Contact-Smoothing---LPS

## Supplementary Material

### Sensitivity analysis for stringency index groupings considering non-holiday periods only

**Figure S1.**
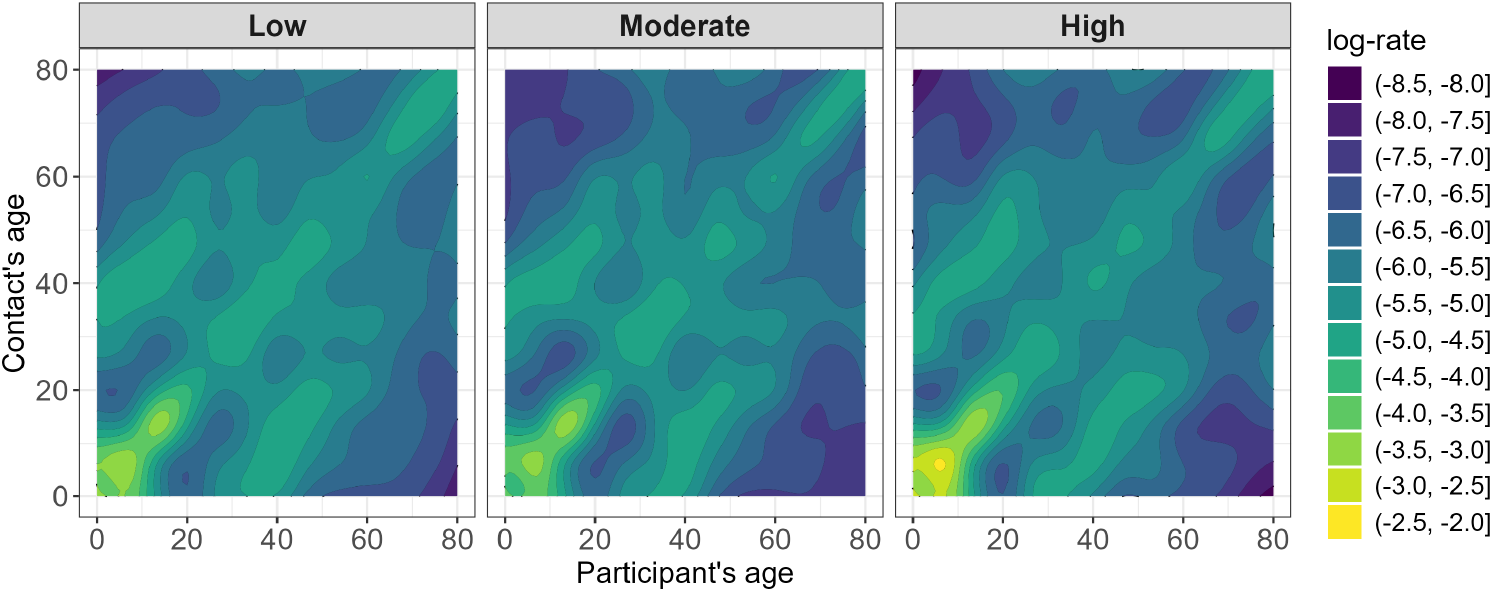
Estimated log-contact rates based on the model with stringency group interaction.

**Figure S2.**
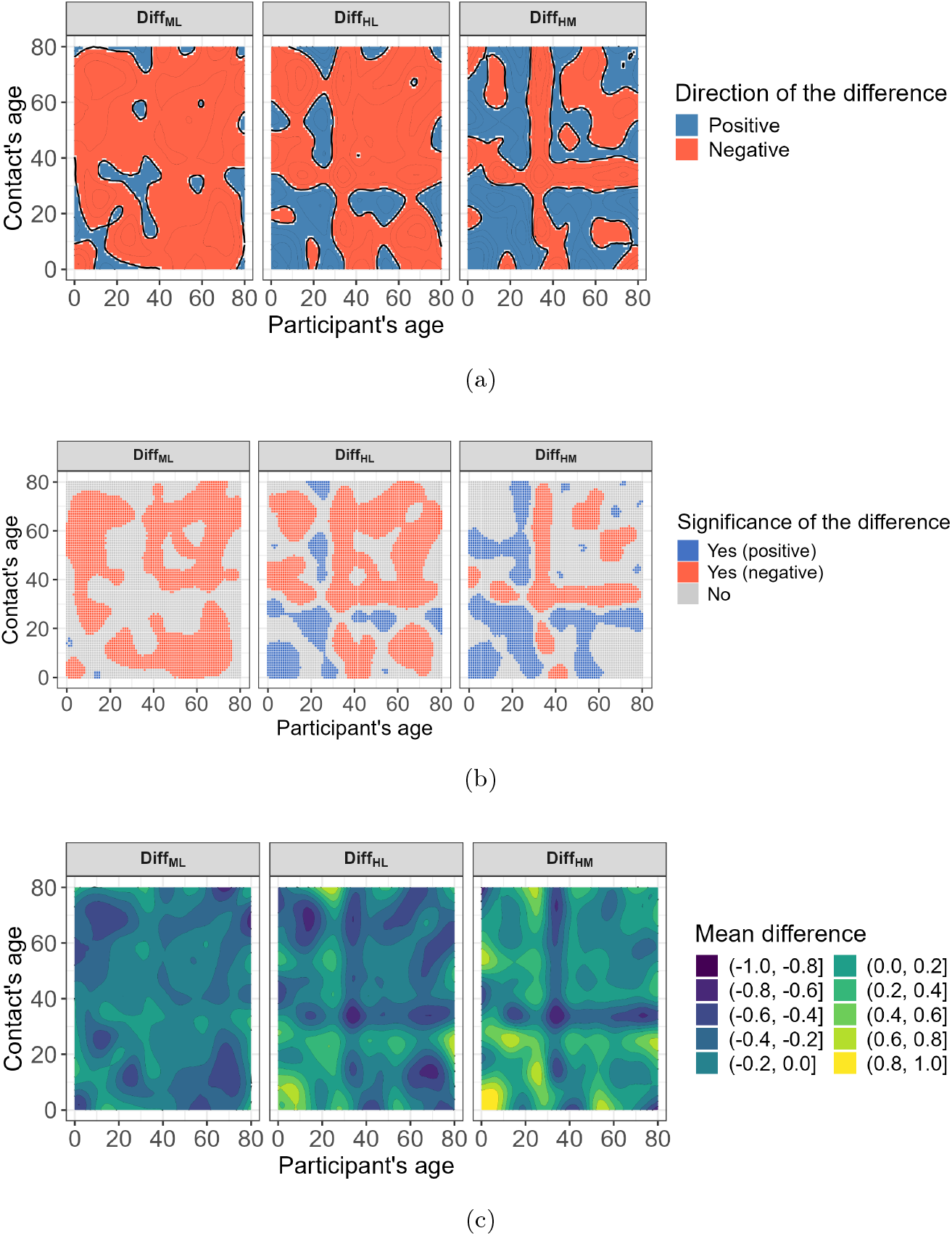
Differences in contact rates between different stringency indices, based on the model with stringency interaction. *Diff*_*ML*_ = Moderate - Low Stringency; *Diff*_*HL*_ = High - Low Stringency; *Diff*_*HM*_ = High - Moderate Stringency. (a) Direction of the difference (*Negative* = fewer contacts compared to low stringency index; *Positive* = more contacts compared to low stringency index); (b) Statistical significance of the difference at a 5% significance level (*Yes (positive)* = Significant and positive direction of the difference; *Yes (negative)* = Significant and negative direction of the difference; *No* = Not significant); (c) Mean difference.

